# Deep Representation Learning of Wrist-Worn Sensor Signals for Latent Motor Abnormality Scoring in Parkinson’s Disease

**DOI:** 10.64898/2026.09.03.26362173

**Authors:** Seyed Mehdi Mohtavipour

## Abstract

Objective quantification of motor abnormality in Parkinson’s disease (PD) remains challenging because conventional clinical assessments are episodic, observer-dependent, and based on coarse ordinal ratings. Wrist-worn wearable sensors provide a scalable opportunity to capture task-specific motor patterns; however, most existing approaches focus on discrete classification rather than deriving continuous latent markers of motor abnormality. In this paper, a multi-stream deep representation network is introduced to derive a latent motor abnormality score using wrist-worn inertial sensor signals. The proposed network includes a local feature extractor based on one-dimensional convolutional layers, a global feature extractor based on Transformer layers, and an embedding layer that maps each task-specific signal into a 64-dimensional embedding vector. A new training procedure is proposed based on a combination of supervised contrastive learning and center loss to cluster the embeddings of PD patients and healthy control (HC) subjects, and to build a latent motor abnormality score based on the average embedding distance from the centroids of PD and HC training embeddings. Across five-fold subject-level cross-validation, the proposed method was assessed for PD/HC classification and achieved an accuracy of 83.10% ± 3.15%, balanced accuracy of 85.89% ± 3.93%, precision of 97.02% ± 2.33%, and ROC-AUC of 0.917. The motor abnormality score showed clear group separation, with healthy controls generally obtaining negative scores and PD subjects obtaining positive scores. Statistical analysis confirmed a significant difference between groups (F = 222.99, p < 0.001), with a substantial effect size (η² = 0.3871). Moreover, exploratory analyses showed that the score tended to increase with higher non-motor symptom burden and longer disease duration, suggesting that the learned latent representation captured disease-related motor abnormalities.

## 1. Introduction

Parkinson’s disease (PD) is a progressive neurodegenerative disorder characterized by a heterogeneous combination of motor and non-motor manifestations. The cardinal motor features of PD include bradykinesia, tremor, rigidity, gait disturbance, and postural imbalance, all of which can substantially affect daily functioning and quality of life [1]. Although PD is primarily diagnosed and monitored through clinical examination, the clinical presentation varies considerably across individuals and disease stages. In particular, motor abnormalities may be subtle in early disease, fluctuate across time, and overlap with other movement disorders that present with parkinsonian features [2]. These challenges motivate the development of objective, quantitative tools that can capture disease-related movement patterns beyond episodic clinical observation. The Movement Disorder Society-Unified Parkinson’s Disease Rating Scale (MDS-UPDRS) remains one of the most widely used clinical instruments for assessing PD symptoms [3]. Its motor examination component provides a structured assessment of bradykinesia, tremor, rigidity, gait, and other motor signs. However, clinical rating scales are inherently limited by their discrete ordinal structure and dependence on expert observation. A single clinical score may compress multiple dimensions of movement impairment into a coarse category, potentially reducing sensitivity to subtle differences in movement quality. Moreover, clinic-based assessments provide only a snapshot of motor function and may not fully reflect the variability of movement performance across tasks, contexts, or repeated measurements. These limitations have stimulated increasing interest in digital measures that quantify motor behavior objectively using wearable and mobile sensor technologies [4].

Wearable inertial sensors are particularly well suited for assessing PD-related motor abnormalities because they can measure acceleration and angular velocity during standardized or free-living activities. Wrist-worn devices are attractive because they are widely available and capable of capturing upper-limb movement patterns that are clinically relevant to PD, including tremor, slowness, rhythm irregularity, reduced movement amplitude, and impaired coordination [5]. Previous studies have demonstrated the potential of wearable sensors and smartwatch-based systems for distinguishing PD from healthy controls, monitoring motor fluctuations, and characterizing task-specific movement impairment [6–8]. Nevertheless, many existing approaches focus primarily on classification performance, treating the model output as a discrete diagnostic decision rather than exploiting the continuous structure learned by the model. This distinction is important because motor impairment in PD is not binary. Individuals may show a continuum of motor abnormality, ranging from healthy-like movement patterns to clearly parkinsonian motor signatures. Furthermore, PD itself is heterogeneous, with differences in symptom dominance, disease duration, medication response, and task performance. A representation-learning framework may therefore provide additional value by learning a latent space in which subjects are organized according to disease-related movement characteristics. Such latent representations can support classification, but they can also be used to define continuous embedding-derived scores that quantify how much an individual’s movement pattern deviates from healthy controls and resembles PD-related motor behavior.

In this work, we propose a deep representation learning framework to capture both local and global features from wrist-worn sensor signals collected during multiple Parkinson’s disease-related motor tasks. The network is designed as a multi-stream architecture, allowing each stream to learn task-specific representations independently before combining the extracted features in the final layers. The local feature extraction layers are intended to capture short-term movement characteristics, such as peaks, cycles, and transient fluctuations, whereas the global feature extraction layers are designed to model temporal relationships among these local patterns across the entire task performed by each subject. The aim of this work is to introduce a continuous latent motor abnormality score. To achieve this, the proposed network uses a supervised contrastive learning approach, in which the final stream-level representations are encouraged to form label-specific clusters in the latent space. This structured embedding space enables the derivation of a continuous abnormality score based on the relative distance of each subject’s embedding from the healthy and PD centroids.

The rest of the paper is organized as follows. Section 2 reviews related work. Section 3 describes the dataset and the proposed network architecture. Section 4 presents the experimental setup and results. Section 5 discusses the main findings, and the final section concludes the paper.

## 2. Related Works

Wearable inertial sensors have increasingly been investigated as objective tools for quantifying motor impairment in PD. Unlike conventional clinical rating scales, which rely on episodic observation and ordinal scoring, wearable sensors can provide continuous, quantitative, and rater-independent measurements of movement. Inertial measurement units, including accelerometers and gyroscopes, have been used to assess tremor, bradykinesia, gait impairment, postural instability, and motor fluctuations in both laboratory and remote settings [9]. For example, authors in [10] investigated different machine learning methods such as deep Convolutional Neural Networks (CNN), bidirectional Long-Short Term Memory (LSTM), or feature-based algorithm on PD subjects gait detection through wrist-worn sensor. They reported that the deep CNN could achieve the best performance on classifying PD subjects with an Area of the Curve (AUC) of 89%. The dataset used in their work included only 18 PD subjects. Another work in [11] proposed two machine learning-based composite models to find the disease progression of PD subjects according to wrist sensor recorded with home monitoring protocol. They evaluated their model on a dataset with 269 PD subjects and could observe a considerable progression on prodromal PD subjects. Ensemble models of different machine learning algorithms with transfer learning technique are introduced in [12] to detect PD disease in a dataset with 55 PD subjects and 30 HC subjects. Authors in this work applied their model on the wrist-worn sensor signal and could achieve about 0.94 of f1-micro metric for PD/HC classification. They also reported that the data from the left-hand sensor were more useful in their classification tasks. A feature-based method is proposed in [13] for PD detection using wrist accelerometer data. In this study, authors utilized statistical features such as signal vector magnitude and movement dispersion to train conventional machine learning methods like Support Vector Machine (SVM) for PD/HC classification in a dataset with 32 HC subjects and 28 PD subjects. They could achieve an average accuracy of 88.5% for the classification task with a reduced set of 20 features. A variability analysis is performed in [14] on wrist-worn sensors for identifying subjects with PD. They showed that the wrist movement variability in HC subjects is greater than subjects with PD. They evaluated their analysis on a data with 29 participants of PD and 29 participants of HC. A simple and reproducible machine learning pipeline is introduced in [15] to detect Parkinson’s disease using a wrist-worn smartwatch signal. They examined several algorithms such as logistic regression, random forest, and CatBoost and showed the logistic regression algorithm could achieve the highest balanced accuracy equal to 79.26% in discriminating HC and PD subjects.

Some other works in the area of PD detection with wrist-worn signal focused on identifying the tremor scoring. In [16] authors proposed a CNN network to process the measured data transformed into 2-dimensional image of frequency domain. They evaluated their approach on a dataset with 92 PD subjects and used UPDRS as references to train the model. They reported an accuracy of 85%. A continuous and unconstrained monitoring algorithm for free-living environments based on supervised machine learning is presented in [17] to identify tremor in PD subjects. Authors in this study categorized time-domain features such as magnitude and variability, frequency-domain features such as power spectral density, and non-linear features such as irregularity to build a model based on SVM machine learning algorithm. They evaluated their approach on a 67-hour database comprising wrist sensor data of 24 PD subjects and demonstrated 90% sensitivity on the test portion of dataset. A data augmentation approach for tremor detection based on wrist-worn signal is presented in [18] where authors utilized generative adversarial network-based method to produce synthesized data and add them to the considered dataset. They reported that with this approach they could boost the obtained accuracy from 74.4% to 80.1% for distinguishing HC and PD subjects.

Existing wearable-sensor studies have demonstrated promising results for PD classification and task-specific motor assessment. However, most of these studies focus on discrete classification rather than continuous latent scoring and further investigation is needed to examine whether representation learning can provide additional information for PD motor assessment. For this purpose, the present work presents a multi-stream deep representation learning framework for wrist-worn sensor signals, using supervised contrastive learning to organize the latent space and derive a continuous embedding-based motor abnormality score.

## 3. Materials and Methods

### 3.1 Dataset, Participants, and Experimental Details

This study used the Parkinson’s Disease Smartwatch (PADS) dataset [19], which contains wrist-worn inertial sensor recordings collected during a standardized neurological motor assessment. The dataset includes recordings from multiple diagnostic groups; however, the present analysis focused on the binary discrimination between Healthy Controls (HC) and individuals with Parkinson’s Disease (PD). After selecting these two groups, the study cohort consisted of 355 subjects, including 79 HC and 276 PD participants. For each subject, wrist movement signals were available from the left and right wrists, recorded using smartwatch inertial sensors at a sampling rate of 100 Hz. Each wrist recording included six kinematic channels: three-axis accelerometer signals and three-axis gyroscope signals. The motor assessment contained multiple task conditions, including resting, postural, and kinematic tasks, such as Relaxed, RelaxedTask, StretchHold, LiftHold, HoldWeight, PointFinger, DrinkGlas, CrossArms, TouchIndex, TouchNose, and Entertainment. Long-duration tasks were divided into two temporal segments to preserve task-specific temporal structure. All signals were resized to a fixed segment length before being used as inputs to the model. The final model therefore used task-specific wrist kinematic streams to learn latent representations for distinguishing PD-like from HC-like motor patterns.

### 3.2 Deep Representation Learning Architecture

In this section, a task-separated multi-stream deep architecture is proposed to learn discriminative wrist-kinematic representations from smartwatch recordings. The overall of this architecture is demonstrated in Fig. 1 where all the 11 input tasks are categorized into 3 different groups such as rest, posture, and kinematic. Relaxed and RelaxedTask are placed in the rest group, StretchHold, LiftHold, and HoldWeight are placed in the posture group, and PointFinger, DrinkGlas, CrossArms, TouchIndex, TouchNose, and Entertainment are placed in the kinematic group. In this architecture, each motor task is processed as an independent stream, allowing the model to learn task-specific latent representations rather than combining all tasks at the input level. For each selected task, signals from both the left and right wrists are used, including three-axis accelerometer and three-axis gyroscope channels. This results in a total of 12 input channels for each stream. The length of each channel is equal to 1024 points for all tasks except for Relaxed, RelaxedTask, and Entertainment where it is 2048, so in this study they are divided into 2 equal 1024 parts. The main part of the proposed deep representation learning is composed of 3 components named as local feature extraction, global feature extraction, and embedding layer with supervised contrastive learning. The local feature extraction part is comprised one-dimensional Convolutional Neural Network (CNN) layers to focus more on attributed such as signal peaks or cycles and every information that are precious at local points. The global feature extraction part is composed of transformer layers to look into finding global patterns like irregularity in each stream data. It is very useful to find relational connections between different time points of stream data which might be placed far from each other. The last part of the proposed architecture is the embedding layer to encode a high dimensional processed channels into a lower dimensional data with contrastive learning supervision. The contrastive learning supervision method used in this study allowed the network to push samples with different labels and pull samples with the same labels. This would result in having a latent space of two separate regions, one for the HC subjects and one for the PD subjects and obtaining a motor abnormality scoring based on the distance from the centroids of these regions. In the next sub-sections, each part of proposed deep representation learning architecture will be described with details.

**Figure 1.**
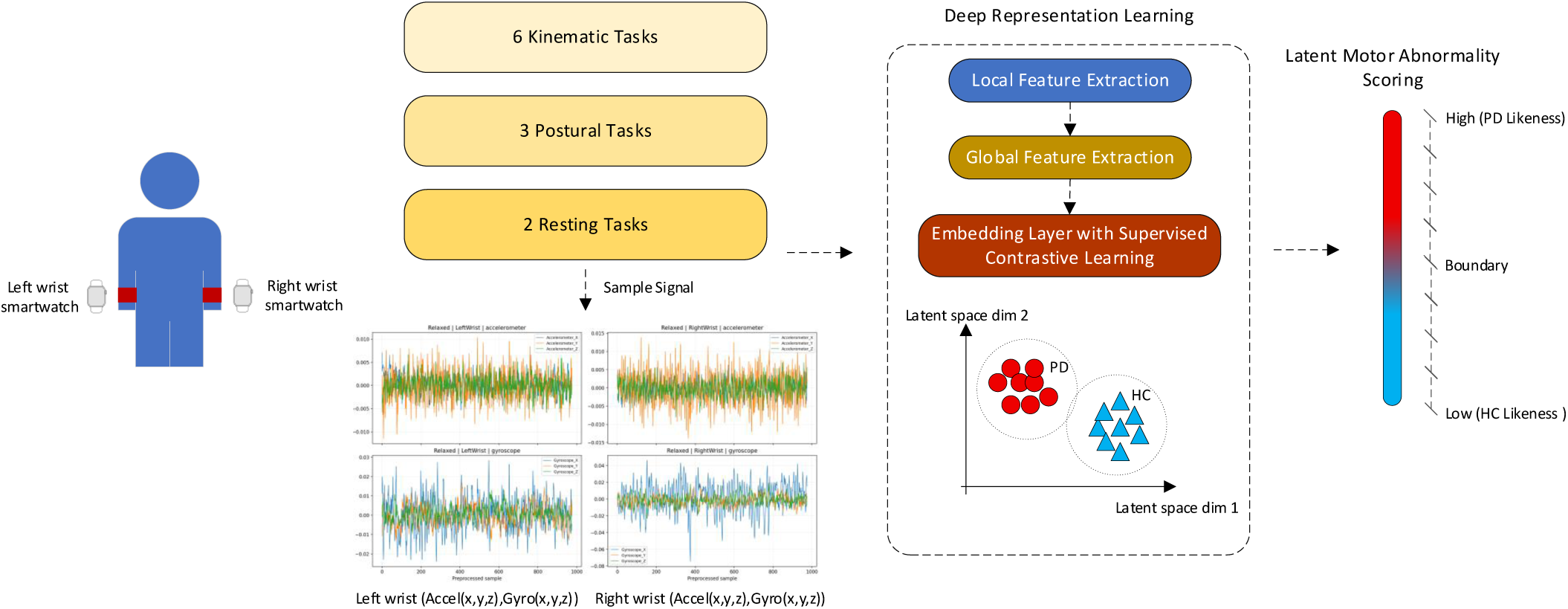
Proposed framework for deriving motor abnormality scoring including 11 sensor signals, deep representation learning network, and score assigning.

#### 3.2.1 Local Feature Extraction

The first part of deep representation learning architecture is local feature extraction to find the local patterns in each input stream and provide a processed and lower dimensional information. For this purpose, one-dimensional CNN layers are employed which are connected sequentially. As can be seen in Fig. 2, for every 11 streams of input data, there are 3 CNN layers to learn independently local features. The first layer mapped the input channels to 8 feature maps using a kernel size of 9 with padding of 4, followed by batch normalization, ReLU activation, and max-pooling with a down-sampling factor of 2. Selecting a larger kernel size at the input level enables the network to learn coarser temporal patterns during the early stages of feature extraction. The convolutional kernel traverses the input signal with the specified stride, generating 8 feature maps corresponding to 8 independently learned filters. After convolution, batch normalization is applied to each feature map according to the following equation.

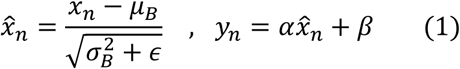

**Figure 2.**
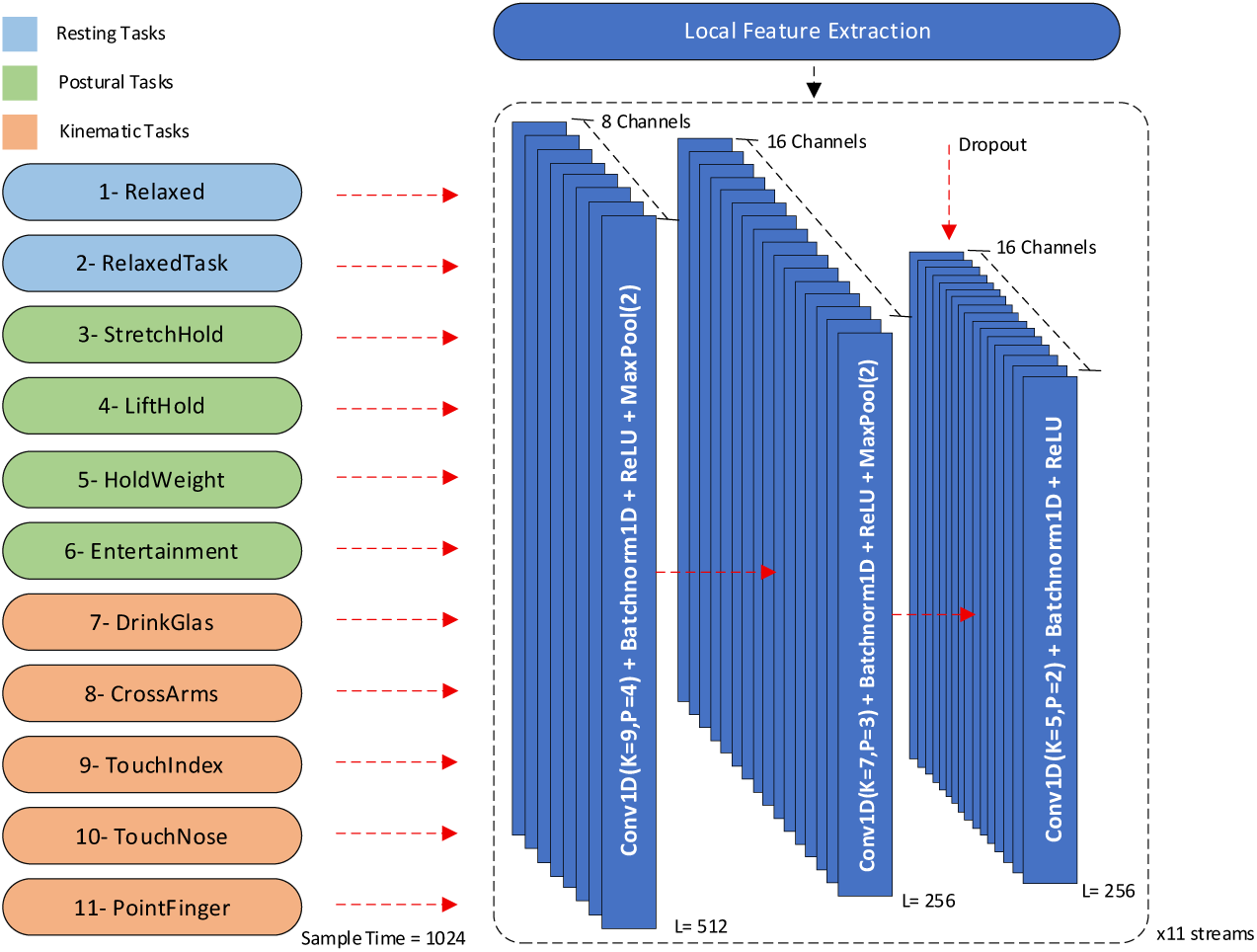
Local feature extraction part of deep representation learning network including convolutional kernels, batch normlaization, ReLU function, max pooling, and dropout.

Where *x_n_* is the convolved data after applying the kernel filter, *μ_B_* and *σ_B_*^2^ are the mean and variance of *x_n_* in each batch, *ε* is a small constant for numerical stability, and *α*, *β* are two learnable scaling and shifting parameters. Batch normalization is applied independently to each channel. The ReLU activation function is applied element-wise to each normalized channel, replacing negative values with zero while leaving positive values unchanged. This activation function is given by the following equation:

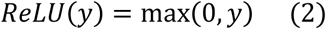

Finally, max pooling examines adjacent elements within each channel and retains the maximum value from each pooling window, thereby reducing the temporal size of the processed feature maps. The second convolutional layer mapped the features to 16 channels using a kernel size of 7 with padding of 3, again followed by batch normalization, ReLU activation, and max-pooling. A third convolutional layer with 16 output channels, kernel size 5, and padding 2 was then applied, followed by batch normalization, ReLU activation, and dropout. The use of progressively smaller convolutional kernels enabled the network to extract local temporal patterns at different scales, while the two max-pooling operations reduced the input sequence length from 1024 to 256 samples before the global feature extraction block. Thus, the CNN module produced a compact temporal representation with 16 feature channels over 256 reduced time points for each task stream.

#### 3.2.2 Global Feature Extraction

After extracting local features through CNN layers, the resulting representation is passed to a transformer block to model longer-range temporal dependencies within each task stream. The final CNN layer produced an output with a shape of ℝ*^B^*^×16×256^where *B* denotes the batch size, 16 is number of feature channels, and 256 is the reduced temporal length. Since the self-attention mechanism is permutation-invariant and does not explicitly encode temporal order, sinusoidal positional encoding was added to the sequence of local feature representations before the Transformer layers. This encoding assigns each time step a deterministic vector composed of sine and cosine functions with different frequencies. By adding these positional vectors to the CNN-extracted features, the Transformer can model not only the relationship between local movement patterns, but also their temporal position within the task. This is particularly important for wrist-worn sensor signals, where the timing and order of peaks, cycles, and transient fluctuations may contain clinically relevant information. The details of the transformer block are demonstrated in Fig. 3 where it can be seen the main building block is the attention component. The attention component considered in this study is a 2-head which each one processes half part of 16 channels. The attention component is composed of three query, key, and value weight matrices with a shape of ℝ^16×16^ which all of them are multiplied with the transposed input features ℝ^256×16^ for each batch denoted as *Y^T^*. The learnable query, key and value of attention component is computed as follows:

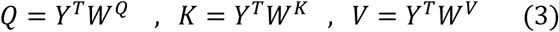

**Figure 3.**
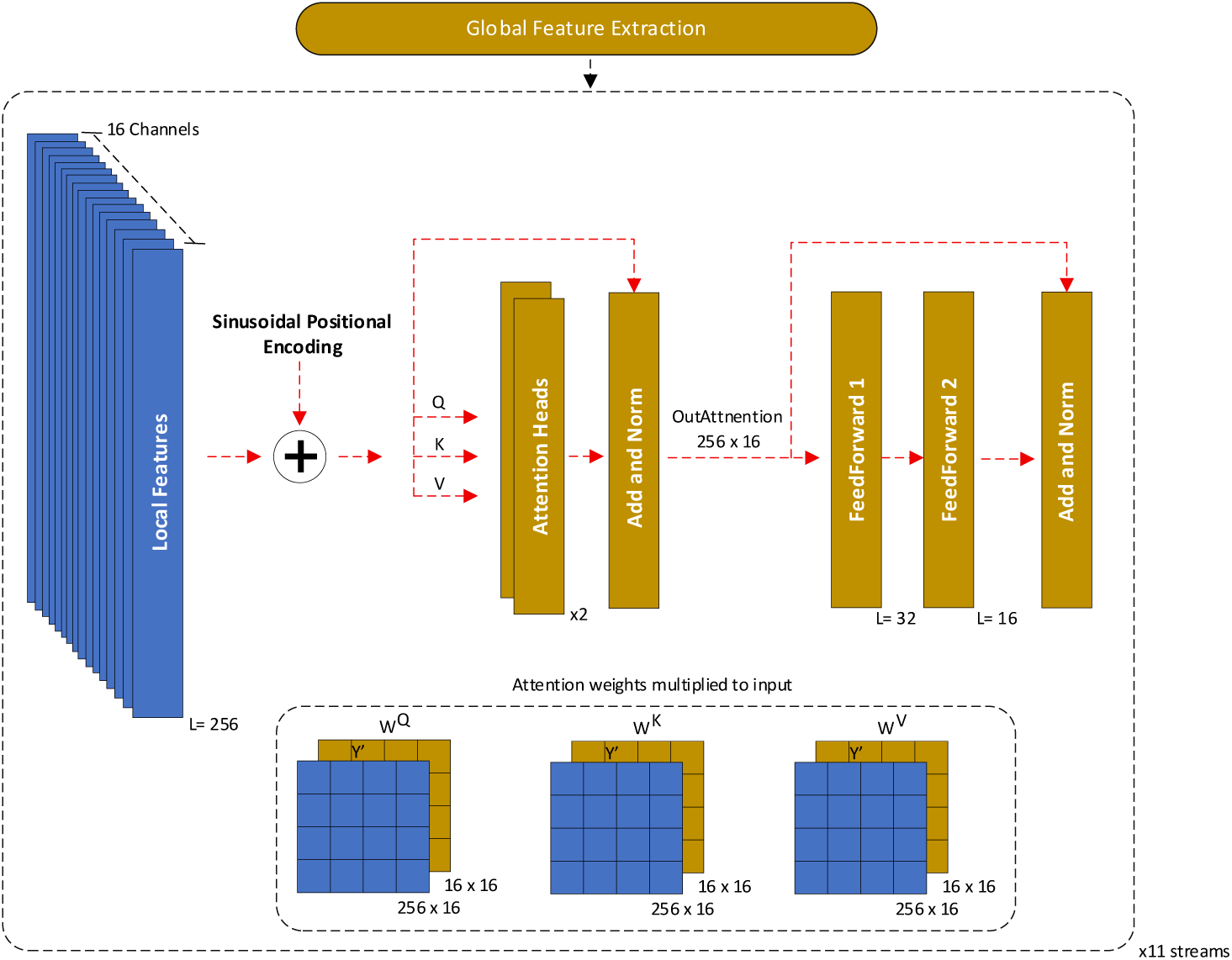
Global feature extraction part of deep representation learning network including attention and feed forward layers.

Where *W^Q^*, *W^K^*, and *W^V^* are the learnable weights of query, key, and value. Each *Q*, *K*, and *V* are split into two equal parts as there are two attention heads in the network and they are denoted by *Q*_ℎ1,2_, *K*_ℎ1,2_, and *V*_ℎ1,2_. The attention scores are then computed by multiplying the query matrix with the transpose of the key matrix. This produces a temporal dependency matrix between all pairs of temporal tokens:

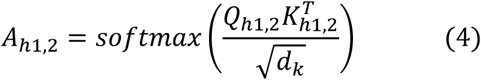

Where softmax function converts the raw attention scores into normalized weight in (0-1) interval, *d_k_* is the dimension of key vector which in this study it is equal to 8, and *T* denotes the matrix transpose function. Finally, the output of each attention head is then obtained by multiplying the attention score matrix with the value matrix as follows:

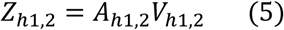

The outputs of the two heads are concatenated and projected back to the original 16-dimensional feature space, after which a residual connection and dropout are applied. Following the attention operation, the feed-forward network consisted of two fully connected layers applied independently to each temporal token. With a transformer dimension of 16 and an MLP ratio of 2.0, the feed-forward sublayer expanded each token representation from 16 to 32 dimensions, and then projected it back from 32 to 16 dimensions. A second residual connection is then used to add the feed-forward output to the attention-enhanced representation. This transformer block therefore preserved the temporal resolution and feature dimensionality of the CNN output, producing a sequence of 256 temporal tokens with 16 features each, while allowing the model to integrate global temporal context across the task-specific wrist signal.

#### 3.2.3 Embedding Layer with Supervised Contrastive Learning

The final part of the proposed deep representation learning architecture is the embedding layer where the processed feature maps produced by local and global extractors are turned into embedding vector and make a latent space such that PD and HC samples are placed far from each other. According to the demonstration in Fig. 4, the output of global feature extraction block which gave feature maps with shape of ℝ^16×256^are passed through an average pooling with two bins to reduce their size into ℝ^16×2^. By flattening the reduced feature maps, a vector with a size of 32 is derived and it makes that possible to construct the last part of embedding layer. This stream-level feature vector is then passed through a task-specific embedding head to map the extracted temporal representation into a discriminative latent space. The embedding head consisted of a fully connected layer that reduced the 32-dimensional feature vector to an 8-dimensional hidden representation, followed by batch normalization, ReLU activation, and dropout. A second fully connected layer is then applied to the hidden representation to turn it into a 64-dimensional embedding vector. The resulting 64-dimensional stream embedding is L2-normalized to constrain the representation to a unit hypersphere. This normalization is used to stabilize contrastive learning and ensure that distances between embeddings are influenced primarily by their relative direction rather than uncontrolled differences in vector magnitude.

**Figure 4.**
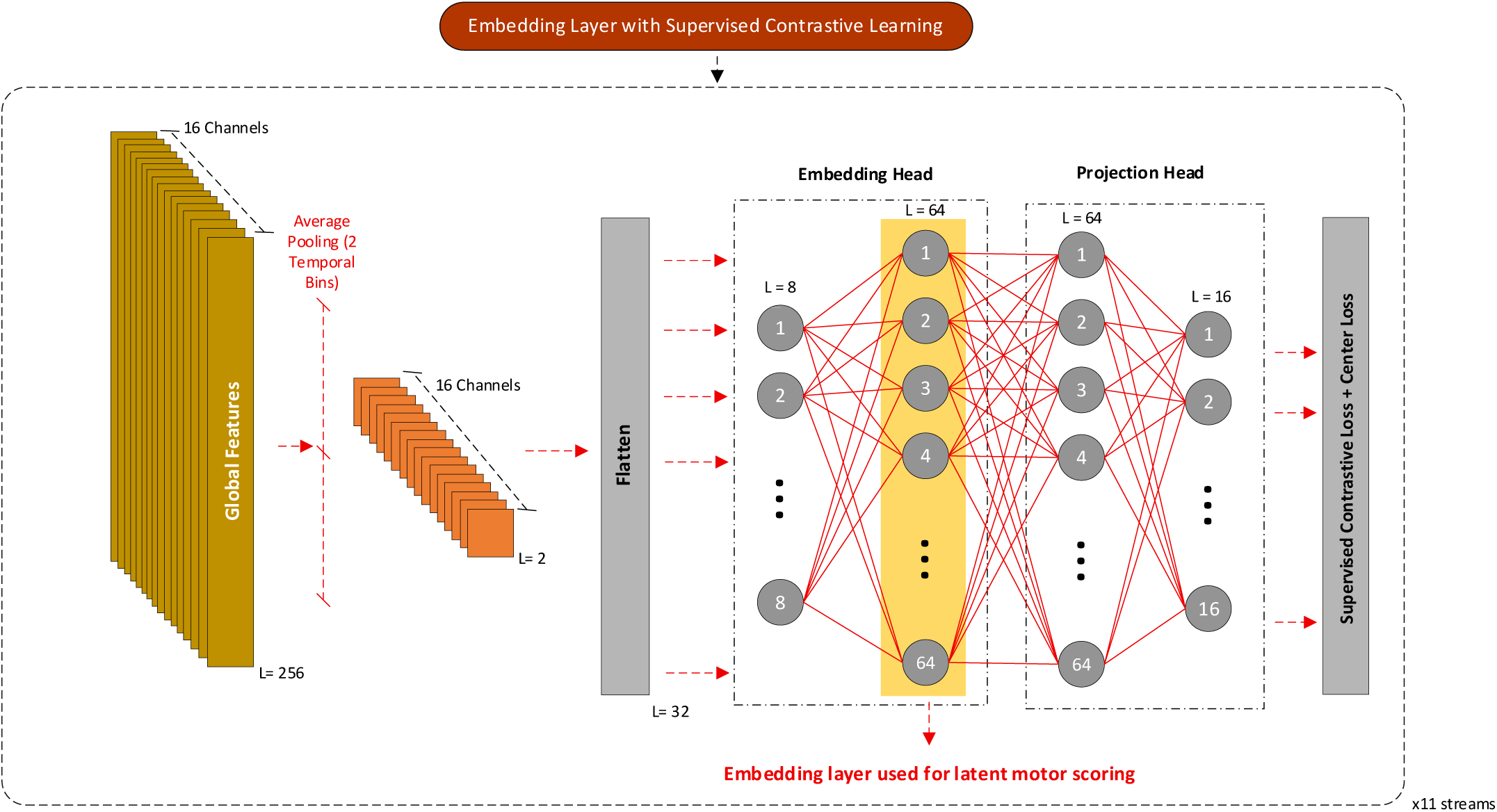
Embedding layer of proposed deep representation learning network, attached with a combination of supervised contrastive learning and center losses.

During training, each normalized stream embedding is further passed through a projection head used only for network learning. This projection head mapped the 64-dimensional embedding to a 16-dimensional contrastive representation through a two-layer fully connected network. Specifically, the projection head first transformed the embedding from 64 to 64 dimensions, applied a ReLU activation, and then projected it from 64 to 16 dimensions, followed by L2 normalization. The projection space is used to compute the supervised contrastive loss, whereas the 64-dimensional vector in the embedding head highlighted in Fig. 4, is retained for latent motor scoring.

The proposed deep architecture is trained according to a Supervised Contrastive Learning (SupCL) loss function introduced in [20] such that the embeddings of PD samples are far from the embeddings of HC samples. Unlike conventional classification losses, supervised contrastive learning explicitly uses class labels to define positive and negative relationships among samples within each batch. Samples belonging to the same class are treated as positive pairs and are encouraged to form compact clusters in the latent space, whereas samples from different classes are treated as negative pairs and are pushed farther apart. As a result, the learned representation becomes more discriminative by increasing intra-class similarity while enhancing inter-class separation. The SupCL loss is defined as follows:

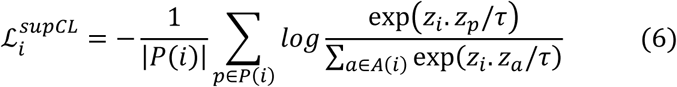

Where *P*(*i*) represents all positive samples in the batch that have the same label as sample *i*, excluding sample *i* itself. *A*(*i*) represents all other samples in the batch, excluding sample *i*, and therefore includes both positive samples from the same label and negative samples from other labels. *z_i_*, *z_p_*, *z_a_* are the latent projection vectors of the anchor sample, positive samples, and all comparison samples, respectively. *τ* is the temperature parameter, which was set to 0.1 in this study. This loss pulls embeddings of samples from the same clinical category closer together and pushes embeddings of samples from different clinical categories farther apart. The SupCL loss for one batch is obtained by averaging the anchor-level losses over all samples:

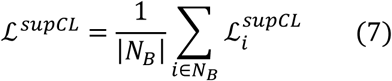

Where *N_B_* is the number of samples in the batch. For making the loss function being more sensitive to separate the embeddings of PD and HC, a second part is added to the loss function as prototype-based center loss. The prototype-based center loss is designed to make the learned embedding space more compact within each class and more separated between classes. Two learnable prototype centers are defined: one for HC, denoted as *c_HC_*, and one for PD, denoted as *c_PD_*. The pulling part of the prototype-based center loss is then defined as:

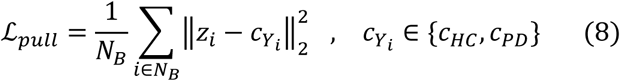

Where *z_i_* is the embedding vector of the anchor sample, and *c_Y_i__* is the prototype center depending on the HC or PD label. This term penalizes large distances between a sample and the prototype center of its true class, thereby promoting intra-class compactness. To improve inter-class separation, a margin-based push term is also introduced. For each sample, the distance to the opposite-class prototype is penalized only when it is smaller than a predefined margin as follows:

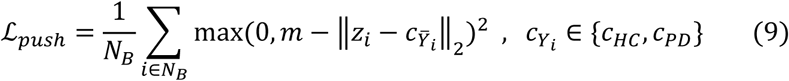

Where *m* is the predefined margin and it is equal to 1.2245 in this study, and *c_Ȳ_i__* is the prototype center of the opposite class. This term discourages embeddings from moving too close to the wrong class prototype. Altogether, the final loss function used in this study is given by the following equation.

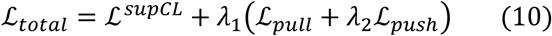

Where *λ*_1_, *λ*_2_ are the coefficients of prototype-based center loss function and in this study, they are considered as 0.2877 and 0.4700, respectively.

### 3.3 Latent Motor Abnormality Scoring

After the completion of network training, the 64-dimensional of embedding vector is utilized to build a latent motor abnormality scoring. For this purpose, first HC and PD class centroids are computed from the training embeddings for each task stream. Let *e_i,s_* be the embedding of subject *i* in stream *s* and *y_i_* ∈ {*HC*, *PD*} be the corresponding class label. The centroid for class *c* in stream *s* is given as follows:

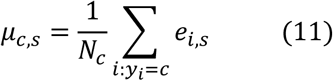

Where *N_c_* is the number of training subjects belonging to class *c*. Thus, each stream has one HC centroid *μ_HC,s_* and one PD centroid *μ_PD,s_*, estimated from training data. For each test subject and every stream, the Euclidean distance between test embedding and each class centroid is computed independently accordingly:

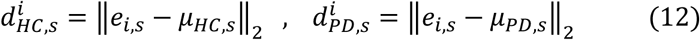

Where *d^i^_HC,s_* and *d^i^_PD,s_* represent the distance of test subject *i* to the to the HC and PD centroids, respectively, in stream *s*. The computed distances are then averaged across all streams to obtain one mean distance to the HC centroids and one mean distance to the PD centroids as follows:

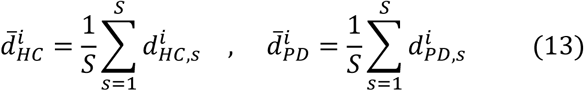

Where *S* is the total number of streams and in this study, it is equal to 11. With the help of these distances, the latent motor abnormality scoring is defined as:

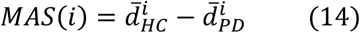

According to this equation, positive values indicate that the subject’s embeddings are, on average, closer to the PD centroids than to the HC centroids, reflecting a more PD-like motor pattern. Conversely, negative values indicate greater proximity to the HC centroids and therefore a more HC-like motor pattern. For classification task, the predicted class is simply assigned based on the smaller averaged distance with the following equation.

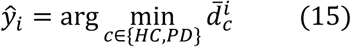

## 4. Experiments and Results

In this section a series of experiments have been conducted to evaluate the effectiveness and performance of proposed deep representation learning approach on capturing motor abnormality in PD subjects. All the experiments of this study are implemented using PyTorch 2.11.0 library on a system with AMD Ryzen 5 5600X 3.70 GHz CPU and GeForce RTX 3060 Ti GPU. The model is trained for 40 epochs using the AdamW optimizer with an initial learning rate of 1.52×10^-3^ and a weight decay of 3.75×10^-3^. A cosine learning-rate scheduler is used to gradually reduce the learning rate to a minimum value of 1×10^-4^. Dropout with a rate of 0.384 is applied during training. Model evaluation is performed only on the held-out test fold in each cross-validation iteration. All experiments are conducted using a fixed random seed of 81. In this section first, a network training assessment is presented to show the performance of the network on HC and PD classification and compared the results with other works. Then an explainability experiment is performed to show which motor tasks have the most contribution on the output of the proposed model. Also, some experiments regarding the relationship between proposed motor abnormality scoring with subjects’ severity or disease duration is presented. Finally, a statistical test is performed to show how the model could separate the PD and HC motor characteristics.

### 4.1 Network Training Assessment

To quantitatively assess the performance of the proposed network, in this section a classification process for the HC and PD subjects is performed based on 5-fold cross validation test and subject data split. For this purpose, all the 355 subjects’ data (including 79 HC and 276 PD subjects) are split into 284 subjects in training and 71 subjects in test for each fold. In the training subjects of each fold, there are 63 HC and 221 PD subjects. Also, in the test subjects of each fold, there are 16 HC and 55 PD subjects. In each fold, one set is selected as test set and others are selected for the training and this moves forward until all subjects are considered once as test subject. Number of samples per batch in the training phase is selected 8 HC and 8 PD subjects. The classification decision is performed based on centroids of HC and PD in each stream during the training phase and then finding the embedding distance of test subjects from them in each stream, averaging across all streams, and assigning a predicted label according to smaller distance from HC and PD centroids. The evaluation metrics for assessing the performance of classification are selected as Accuracy, Balanced Accuracy, Precision, Recall, and F1-score which are computed according to True Positive (TP), True Negative (TN), False Positive (FP), and False Negative (FN) rates. True Positives (TP) denote samples correctly assigned to the PD class, while False Positives (FP) denote samples incorrectly assigned to PD class despite belonging to HC. False Negatives (FN) denote samples belonging to the PD class but incorrectly assigned to HC class. True Negatives (TN) denote samples from HC class that are correctly not assigned to the PD class. Accuracy and Balanced Accuracy are defined as follows:

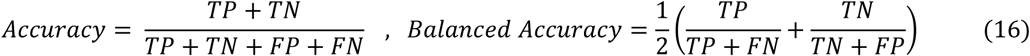

The Balanced Accuracy is a more appropriate metric in this evaluation as the dataset is imbalanced in terms of numbers PD and HC subjects. Precision and Recall metrics are also defined as follows:

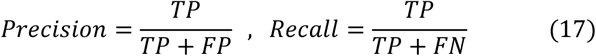

And the F1-score is equal to:

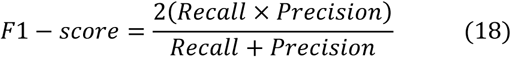

Fig. 5 presents the confusion matrix of all predicted samples appeared in the test subset of data. Since there are 5 folds in the data split, the results for the prediction of held-out test subset in each fold are summed to produce the TP, TN, FP, and FN. As can be seen, from 79 subjects in the HC set, the proposed network could predict most of them correctly with only 7 misclassified subjects. In the PD set, the number of incorrectly predicted is higher with 53 subject error which results in an 81% correction rate. Higher number of errors in PD test can be justified according to the diverse severity and there might be PD subjects with lower severity performing tasks much similar to HC subjects. Table 1 shows the results according to the evaluation metrics across the five-fold cross-validation. The proposed model achieved an average accuracy of 83.10% ± 3.15% and an average balanced accuracy of 85.89% ± 3.93%, indicating stable performance despite the class imbalance between HC and PD subjects. The model showed particularly high precision for the PD class, with an average precision of 97.02% ± 2.33%, suggesting that samples predicted as PD were rarely misclassified HC subjects. The average PD recall was 80.79% ± 3.83%, indicating that most PD subjects were correctly detected, although a subset was still classified as HC. The resulting average PD F1-score was 88.11% ± 2.40%, reflecting a strong balance between precision and recall. Across individual folds, balanced accuracy ranged from 81.93% to 90.00%, while PD precision remained consistently high, reaching 100% in fold 4.

**Figure 5.**
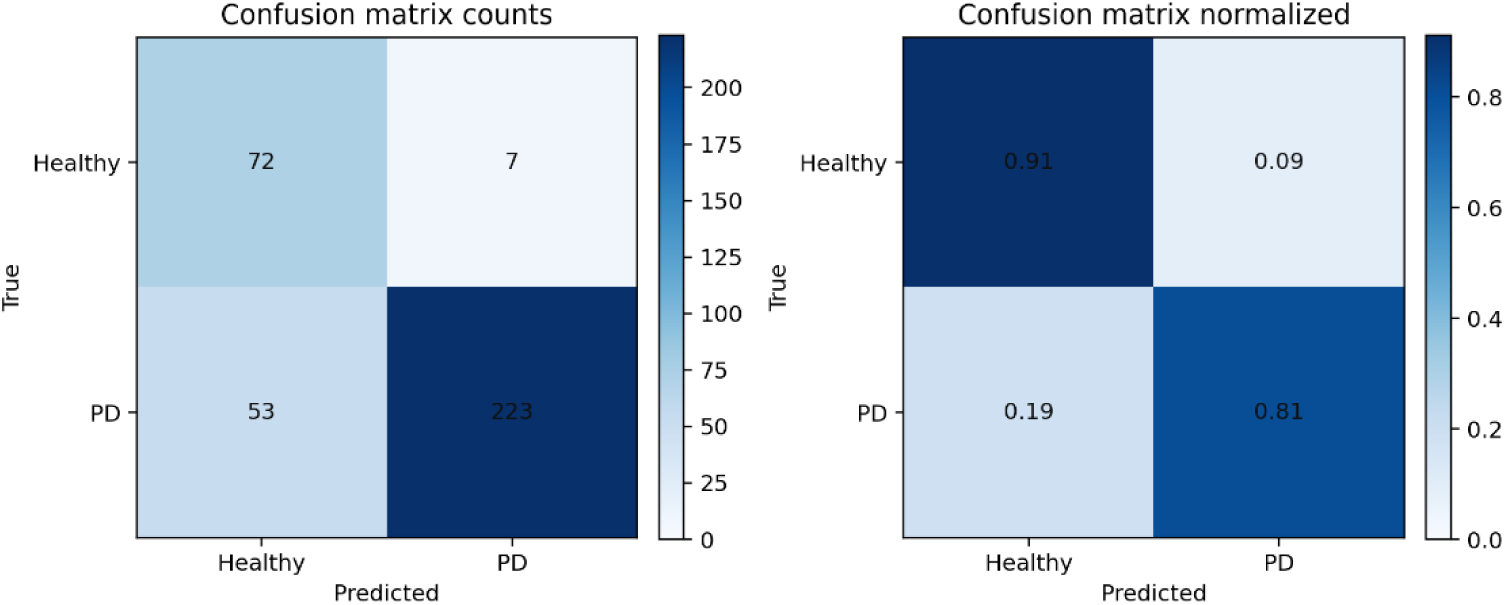
Confusion matrix of test prediction summation appeared in all 5-fold of cross validation.

**Table 1.** Cross validation classification results for the proposed deep representation learning network.

| Fold Number | Accuracy | Balanced Accuracy | Precision | Recall | F1-score |
| --- | --- | --- | --- | --- | --- |
| #1 | 87.32% | 89.60% | 97.92% | 85.45% | 91.29% |
| #2 | 81.69% | 85.97% | 97.73% | 78.18% | 86.87% |
| #3 | 78.87% | 81.93% | 95.45% | 76.36% | 84.85% |
| #4 | 84.51% | 90.00% | 100% | 80.00% | 88.89% |
| #5 | 83.10% | 81.96% | 94.00% | 83.93% | 88.68% |
| **Average** | $83.10\% \pm 3.15\%$ | $85.89\% \pm 3.93\%$ | $97.02\% \pm 2.33\%$ | $80.79\% \pm 3.83\%$ | $88.11\% \pm 2.40\%$ |

Another important metric that can be used to evaluate the discriminative ability of the proposed model across different classification thresholds is the receiver operating characteristic (ROC) curve. The ROC curve plots the true positive rate across the false positive rate. Unlike accuracy, which depends on a fixed decision threshold, the ROC curve summarizes model performance over all possible thresholds. The area under the ROC curve (AUC) provides a threshold-independent measure of separability, where a value of 1.0 indicates perfect discrimination and a value of 0.5 indicates random classification. Fig. 6 shows the results of ROC for all held-out test predictions of 5 folds and also the aggregated held-out test predictions of all folds. Area Under Curve (AUC) of ROC for aggregated test predictions is computed 0.917, indicating excellent overall classification performance and high separability between the two groups. The curve rises steeply toward the upper-left region of the plot, showing that the model can achieve a high true positive rate while maintaining a relatively low false positive rate across a wide range of decision thresholds. In addition, the ROC curves from the individual folds follow a similar trend and remain consistently above the diagonal reference line, suggesting that the model performance is stable across cross-validation folds and substantially better than random classification.

**Figure 6.**
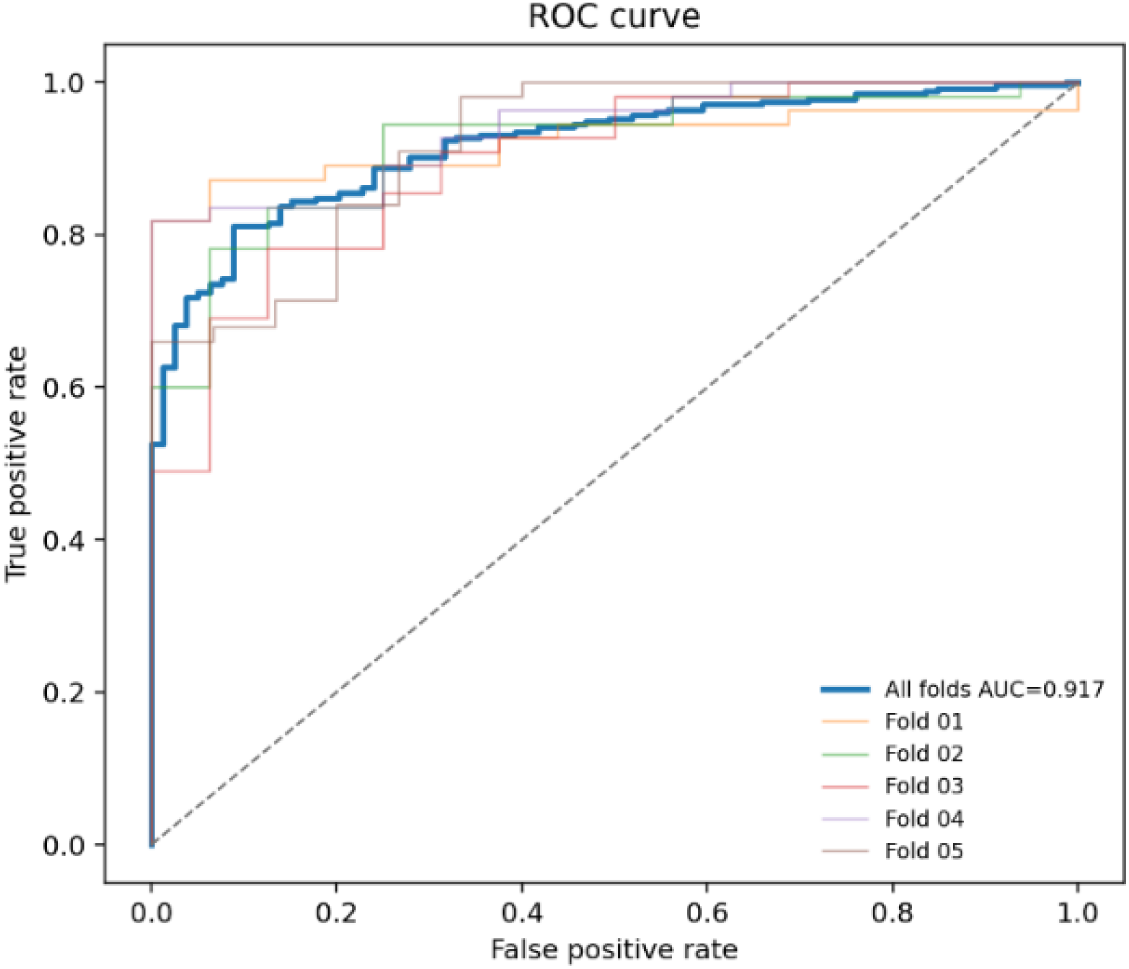
ROC and AUC results across all 5-fold cross validation.

### 4.2 Classification Comparison with Other Works

Table 2 compares the proposed method with previous studies on the PADS dataset. The proposed approach achieved a balanced accuracy of 85.89% ± 3.93%, outperforming the reported PADS baseline (78.99% ± 7.66%) and the classical machine learning approach (79.26% ± 5.50%) in terms of class-balanced performance. Although the classical machine learning method reported a higher overall accuracy (87.32% ± 1.99%) than this work (83.10% ± 3.15%), its lower balanced accuracy suggests reduced robustness under class imbalance. It should also be noted that the PADS baseline used different network setups and configurations across folds, which may limit direct comparability with the present work, where a consistent model configuration was used across the cross-validation procedure. The GAN-synthesized tremor approach reported an accuracy of 80.10% ± 5.70%, but balanced accuracy was not available, limiting direct comparison. Overall, these results suggest that the proposed method provides more balanced and reliable classification performance across classes.

**Table 2.** Classification comparison with other works in terms of accuracy and balanced accuracy.

| Method | Accuracy | Balanced Accuracy |
| --- | --- | --- |
| **PADS baseline [19]** | N/A | $78.99\% \pm 7.66\%$ |
| **GAN synthesized Tremor [18]** | $80.10\% \pm 5.70\%$ | N/A |
| **Classical ML [15]** | $87.32\% \pm 1.99\%$ | $79.26\% \pm 5.50\%$ |
| **This work** | $83.10\% \pm 3.15\%$ | $85.89\% \pm 3.93\%$ |

### 4.3 Explainability Assessment with Input Gradient Saliency

To further interpret the trained model and identify which parts of the wrist kinematic recordings contributed most strongly to the classification decision, a gradient-based saliency analysis is performed. For each held-out test sample, the gradient of the motor abnormality score with respect to the input signal is computed. Then the absolute gradient magnitude is averaged across time points, sensor axes, and both wrists within each task-specific stream, yielding one saliency value per stream for each held-out test sample. Higher saliency values indicate that small changes in the corresponding signal region would have a stronger influence on the model’s abnormality score. This analysis provides an interpretable view of the task and wrist channels that are most influential in distinguishing PD-like from HC-like motor patterns. Fig. 7 shows the mean absolute saliency for the proposed motor abnormality scoring network. It revealed that the model relied most strongly on posture- and load-related tasks. The highest mean absolute saliency values are observed for the StretchHold and LiftHold streams, suggesting that these tasks contributed most substantially to the model output. This indicates that sustained posture and holding tasks may contain particularly informative wrist-sensor patterns for identifying PD-related motor abnormality, potentially reflecting impaired steadiness, tremor-related fluctuations, or altered motor control during sustained movements. Relaxed and PointFinger streams also showed moderate saliency, indicating that both resting-state and active movement information contributed to the learned representation. In contrast, CrossArms, TouchIndex, DrinkGlas, TouchNose, and Entertainment showed lower saliency values, suggesting a smaller average influence on the model decision. Overall, this analysis supports the relevance of activity-specific stream modeling and indicates that the proposed network does not rely equally on all smartwatch tasks, but instead assigns greater sensitivity to streams containing more discriminative motor information.

**Figure 7.**
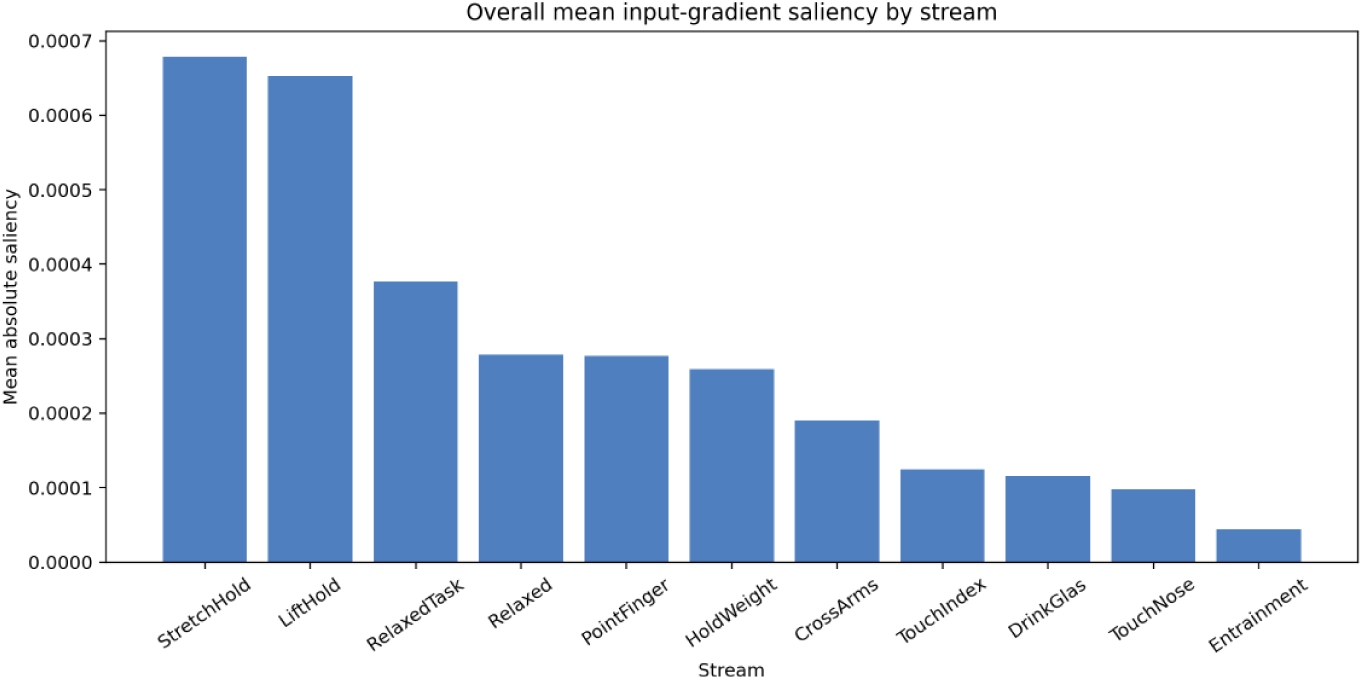
Results for mean gradient saliency of all streams in the proposed deep representation learning network.

### 4.4 Motor Abnormality Score Distribution across HC and PD labels

In this experiment, the proposed motor abnormality scoring is evaluated across the ground truth labels of all subjects in the dataset. To obtain the motor abnormality score of each subject, first the embedding vectors of all streams in the corresponding held-out test fold are computed. For each stream, the Euclidean distance between the test embedding and the HC and PD centroid vectors is computed. These stream-wise distances are then averaged across all streams, yielding one mean distance to the HC centroids and one mean distance to the PD centroids. The motor abnormality score is equal to the subtraction of averaged distance to HC centroid and PD centroid, respectively. This metric reflects the extent to which a test subject’s embedding is closer to HC-like or PD-like patterns. The more positive the motor abnormality score is, the more PD likeness is observed. On the contrary, the more negative the motor abnormality score is, the more HC likeness is observed. Fig. 8 shows the density distribution of HC and PD population across the motor abnormality scoring. As can be seen, most healthy samples obtained negative scores, indicating that their latent representations were closer to the healthy centroid, whereas most PD samples obtained positive scores, indicating greater proximity to the PD centroid. The decision boundary at zero therefore provides an interpretable separation criterion based on relative centroid distances in the learned latent space. Although some overlap is observed around the boundary, reflecting ambiguous or borderline samples, the overall shift of the PD distribution toward positive values and the healthy distribution toward negative values supports the ability of the learned representation to encode PD-related motor abnormality. This result further motivates the use of the latent score as a continuous motor abnormality index rather than only a binary classification output.

**Figure 8.**
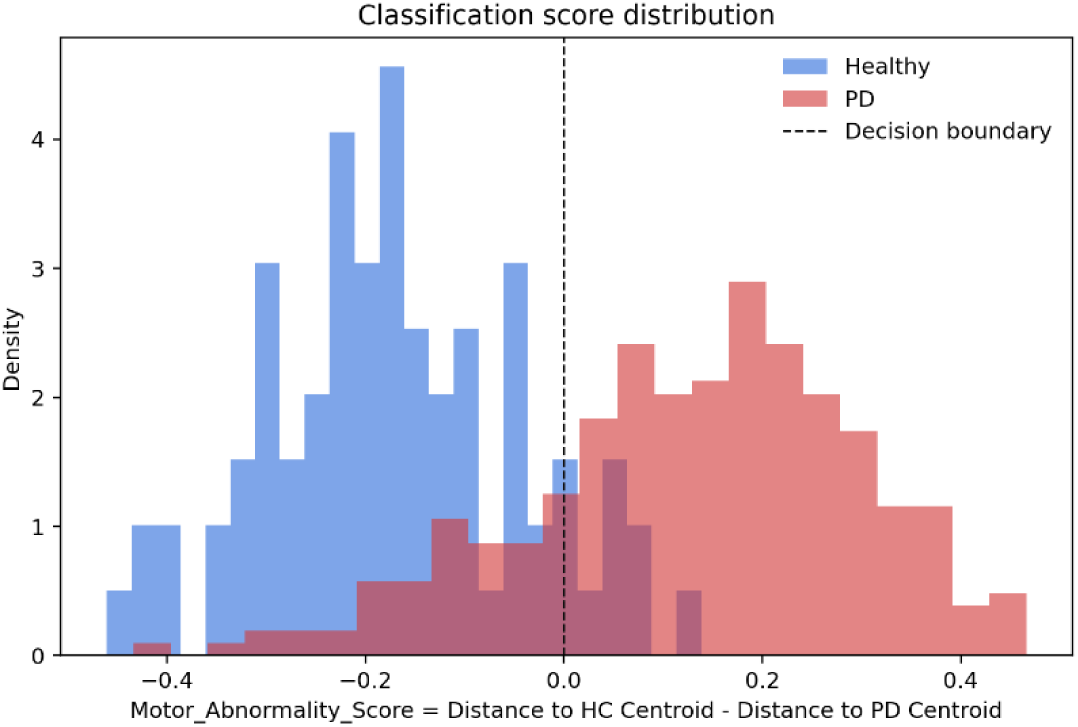
Distribution of motor abnormality scores for HC and PD subjects across all held-out test folds.

### 4.5 Association Between Motor Abnormality Score and NMS Burden

To explore the clinical relevance of the proposed score, it is necessary to compare the scores with clinical severity ratings for PD subjects. Since clinical severity ratings are not available in this dataset, one possible approach is to use the Non-Motor Symptoms (NMS) questionnaire provided in the dataset as an approximation of clinical severity. The NMS questionnaire includes 25 questions covering a wide range of non-motor symptoms, such as hallucinations, depression, and sleep problems, with binary true/false responses. Therefore, in this paper, we assume that subjects with a higher number of positive responses on the NMS questionnaire have higher clinical severity. For this purpose, the number of positive NMS responses was categorized into five bins: (0–4), (5–9), (10–14), (15–19), and (20–24). The proposed abnormality score is then plotted across these NMS response bins for each subject. The relationship between the proposed latent motor abnormality score and the binned NMS questionnaire total is shown in Fig. 9 healthy controls were mostly concentrated in the lowest NMS bin and generally showed negative abnormality scores, indicating proximity to the healthy region of the latent space. In contrast, PD subjects were distributed across all NMS bins and predominantly showed positive abnormality scores. The bin-level mean increased from the lowest NMS category to the intermediate categories, suggesting that higher non-motor symptom burden is generally associated with increased latent motor abnormality. However, this trend was not strictly monotonic, particularly in the highest NMS bin, where larger variability and fewer samples may affect the mean estimate. These findings suggest that the proposed score captures PD-related motor abnormality and shows an exploratory association with NMS burden, while also highlighting that NMS should be interpreted only as an indirect proxy rather than a direct clinical motor severity measure.

**Figure 9.**
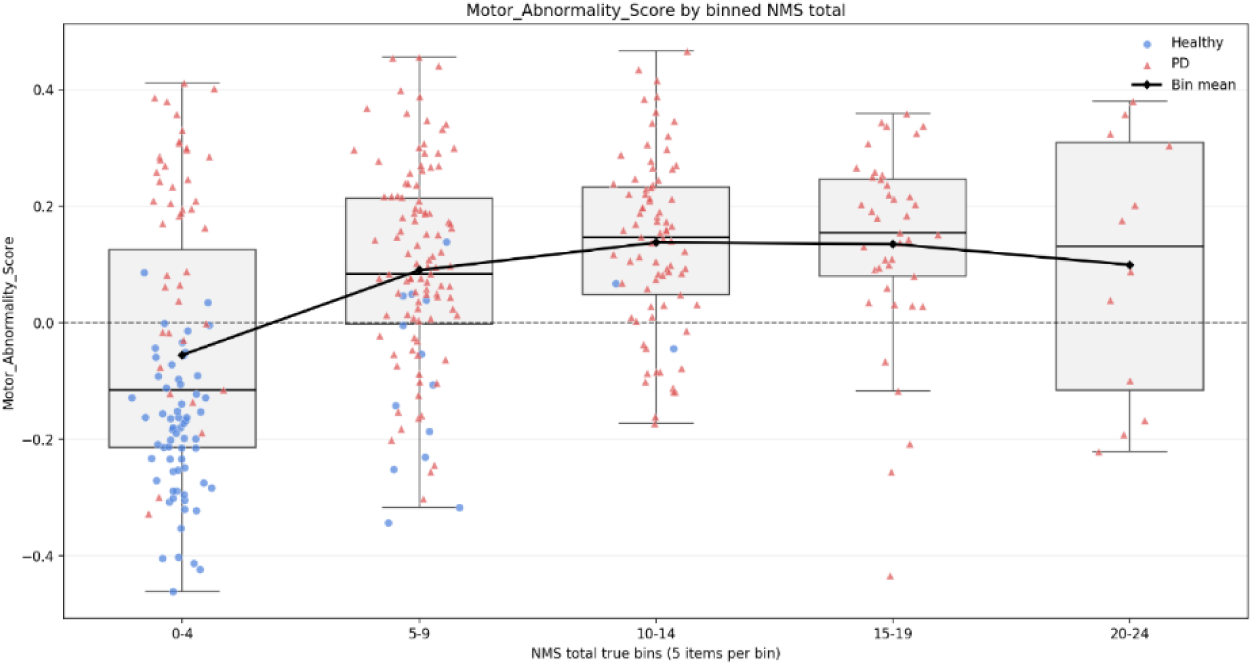
Distribution of the proposed motor abnormality score across five bins of positive responses on the Non-Motor Symptoms (NMS) questionnaire.

### 4.6 Association Between Motor Abnormality Score and Disease Duration

Another exploratory analysis of the proposed motor abnormality score is evaluating it across the disease duration. This analysis explores whether the proposed score shows a trend with longer disease duration. For this purpose, disease duration is grouped into five-year intervals, including (0–4), (5–9), (10–14), (15–19), and more than 20 years. For each bin, the distribution of the motor abnormality score is visualized using boxplots, with individual subject scores overlaid as scatter points. As shown in Fig. 10, the motor abnormality score remained predominantly above the zero-reference line across all disease-duration bins, indicating that most PD subjects were positioned closer to the PD centroid than to the healthy centroid in the learned latent space. A gradual increase in the bin-level mean is observed from the shortest disease-duration group toward the longer-duration groups. This suggests that subjects with longer disease duration tend to show higher latent motor abnormality scores, reflecting that the embedding-derived score captures disease-related motor burden. However, substantial within-bin variability is observed, particularly in the shorter disease-duration groups. The distributions across bins also showed considerable overlap, indicating that disease duration alone does not fully explain the variability in the proposed score. This is expected, as motor impairment in PD is influenced by multiple factors, including disease phenotype, medication state, symptom dominance, compensatory movement strategies, and individual differences in task performance. In addition, the highest disease-duration bins contained fewer subjects, which may reduce the stability of the estimated bin-level means. Overall, these results provide exploratory evidence that the proposed latent motor abnormality score is associated with disease duration in PD subjects. The observed trend suggests that the score may reflect clinically meaningful disease-related motor changes.

**Figure 10.**
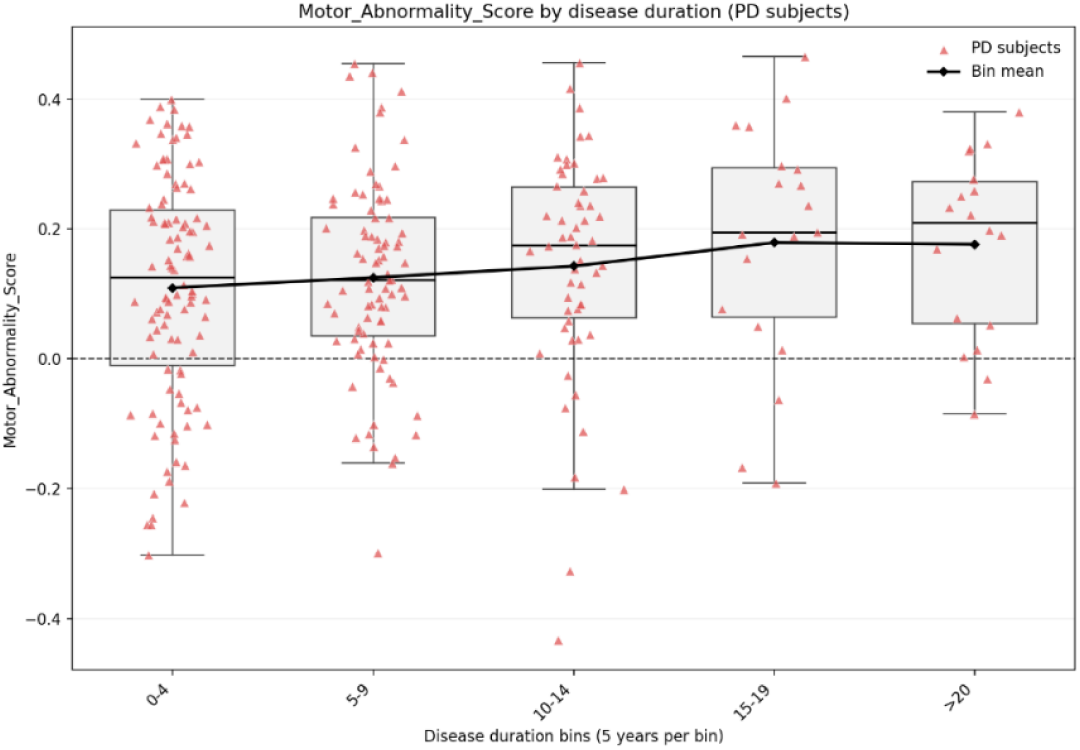
Distribution of the proposed motor abnormality score across five bins of disease duration.

### 4.7 Sensitivity to PD Subtype

In this section, sensitivity of the proposed score to different PD subtypes is evaluated. Fig. 11 shows the distribution of the proposed motor abnormality score across PD subtypes derived from the available disease-comment information. Overall, all PD subtype groups show predominantly positive abnormality scores, indicating that subjects from each subtype are generally positioned closer to the PD region than to the healthy region in the learned latent space. The median score appears slightly higher in the mixed and other PD comment groups compared with the akinetic-rigid and unknown groups, while the tremor-dominant group shows a broad distribution with substantial variability. However, there is considerable overlap among all subtype distributions, suggesting that the proposed score captures a general PD-related motor abnormality rather than a subtype-specific pattern.

**Figure 11.**
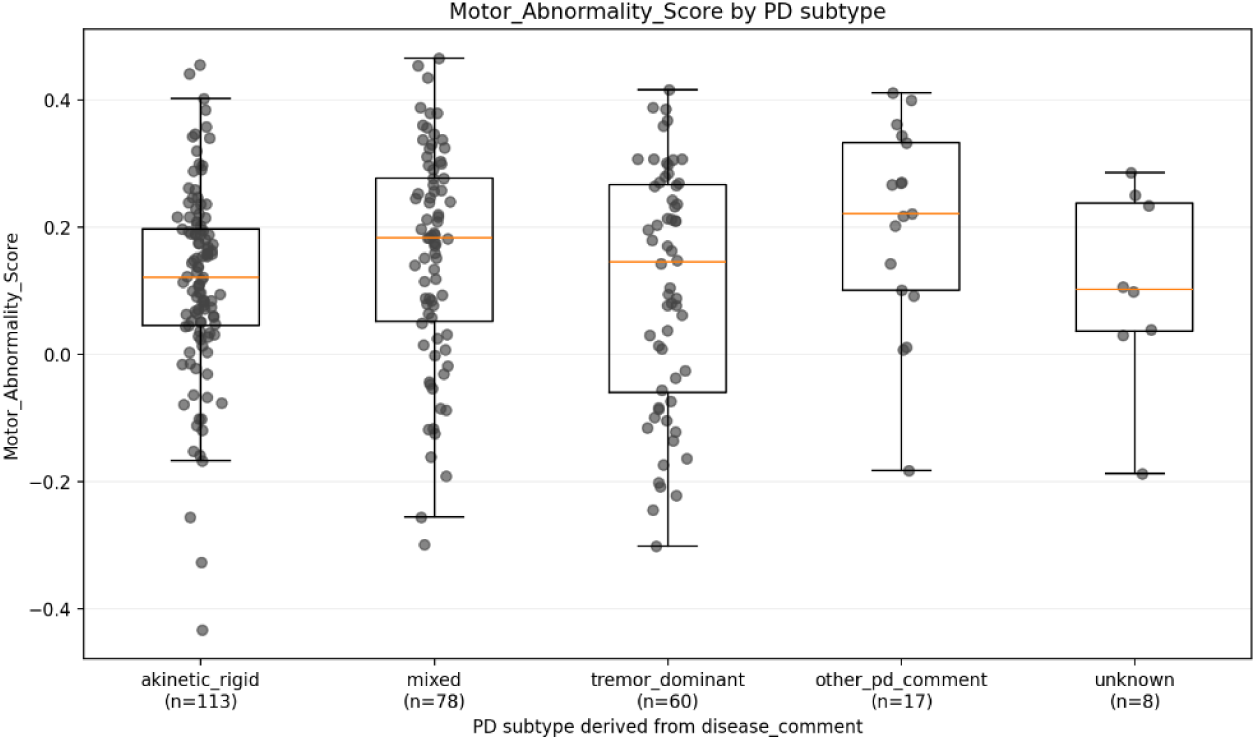
Distribution of the proposed motor abnormality score across five PD subtype groups.

### 4.8 Statistical Separation of HC and PD

In this section an Analysis of Variance (ANOVA) is performed to show statistically significant differences of the measures for HC and PD categories. Table 3 shows the ANOVA test result and it can be seen that there is a significant difference in the proposed motor abnormality score between groups, with an F-statistic of 222.99 and a p-value of < 0.001. The effect size is 0.3871 with a 95% confidence interval of (0.3164–0.4604), indicating a substantial group effect. The mean score is negative for healthy controls (−0.1720) and positive for PD subjects (0.1298), suggesting that healthy subjects are generally closer to the healthy centroid, whereas PD subjects are closer to the PD centroid in the learned latent space. These findings provide statistical support for the group-level separation encoded by the proposed score and indicate that the embedding-derived score captures meaningful PD-related motor differences.

**Table 3.** ANOVA test result for motor abnormality scoring across HC and PD subjects.

| Feature | F-statistic | p-value | Effect size ($\eta^2$) | Confidence interval | HC mean | PD mean |
| --- | --- | --- | --- | --- | --- | --- |
| Motor abnormality score | 222.99 | $< 0.001$ | 0.3871 | (0.3164-0.4604) | -0.1720 | 0.1298 |

## 5. Discussion

In this study, a multi-stream deep representation learning framework is proposed to derive a latent motor abnormality score from wrist-worn inertial sensor signals in Parkinson’s disease. Unlike conventional classification-based approaches, the proposed method is designed to learn a structured embedding space in which healthy control and PD subjects are organized according to their wrist-movement characteristics. By combining one-dimensional convolutional layers for local temporal feature extraction, Transformer-based layers for global temporal modeling, and supervised contrastive learning with prototype-based center loss, the model was encouraged to learn compact within-class representations while increasing the separation between HC-like and PD-like motor patterns. This design enabled the derivation of a continuous motor abnormality score based on the relative distance of each subject’s embedding to HC and PD centroids. The proposed network was assessed through five-fold subject-level cross-validation, and could achieve an average accuracy of 83.10%, balanced accuracy of 85.89%, and an AUC of 0.917, indicating strong separation between HC-like and PD-like motor patterns. The balanced accuracy is particularly important given the class imbalance between HC and PD subjects, suggesting that the model did not simply benefit from the larger number of PD samples. However, the difference between the high precision and lower recall indicates that the model was more conservative in assigning subjects to the PD class, with some PD subjects being classified as HC. This may reflect the heterogeneity of PD and the possibility that mildly affected subjects can show wrist-movement patterns close to those of healthy controls. The proposed latent motor abnormality score provided a continuous interpretation of the learned embedding space. HC subjects generally obtained negative scores, indicating greater proximity to the HC centroid, whereas PD subjects generally obtained positive scores, indicating greater proximity to the PD centroid. The ANOVA results confirmed a significant group difference with a substantial effect size, supporting the discriminative validity of the score.

The exploratory analyses with NMS burden and disease duration provide additional but indirect evidence for the clinical relevance of the proposed latent motor abnormality score. The score showed a tendency to increase in subjects with a higher number of positive NMS responses and in subjects with longer disease duration, suggesting that the learned latent representation may capture broader disease-related motor burden beyond the binary separation of HC and PD groups. This trend suggests that subjects with more PD-like embeddings may also have a higher overall disease burden. Motor manifestations in PD can vary substantially across individuals due to differences in phenotype, medication state, compensatory strategies, and progression rate. Therefore, these analyses should be considered exploratory and hypothesis-generating, supporting the potential clinical relevance of the score while highlighting the need for future validation against established motor severity scales such as MDS-UPDRS Part III.

The PD subtype analysis showed predominantly positive abnormality scores across subtype groups, indicating that the score captures a general PD-related motor signature. However, the substantial overlap between subtype distributions suggests that the current representation is not strongly subtype-specific. This is expected because subtype labels were not used during training, and the model was optimized for HC/PD separation rather than differentiation among PD phenotypes. Future subtype-aware or multi-task learning approaches may be required to determine whether wrist-worn sensor embeddings can capture more specific motor phenotypes such as tremor-dominant or akinetic-rigid patterns.

The input-gradient saliency analysis indicated that the model was most sensitive to posture- and load-related tasks, particularly StretchHold and LiftHold. This suggests that sustained upper-limb control tasks may contain important information for distinguishing PD-like from HC-like motor patterns, potentially reflecting impaired steadiness, tremor-related fluctuations, or altered motor control during sustained postures. Overall, the findings support the value of task-specific multi-stream representation learning and show that smartwatch-derived embeddings can provide both robust classification performance and a continuous motor abnormality score.

## 6. Conclusion

This paper presented a multi-stream deep representation learning approach for deriving a latent motor abnormality score from wrist-worn inertial sensor signals. The proposed framework processed multiple standardized motor tasks through task-specific streams and organized the learned embeddings into HC-like and PD-like regions using supervised contrastive learning and center loss. By defining the score based on the relative distance to HC and PD centroids, the model provided both a classification decision and a continuous measure of PD-like motor abnormality. The experimental results demonstrated that the proposed method can distinguish HC and PD subjects with strong class-balanced performance. Beyond classification, the derived score provided a representation of each subject’s position in the learned latent motor space. Exploratory analyses suggested that this score is related to disease-associated characteristics, including non-motor symptom burden and disease duration, while saliency analysis highlighted the importance of posture- and load-related tasks. These findings indicate that smartwatch-based representation learning may provide a scalable and objective approach for quantifying PD-related motor abnormality. Longitudinal study and tracking the disease progression according to changes of proposed abnormality scoring over time would be the future work of this study.

## Data Availability

All data used in the present study are publicly available online through the Parkinson's Disease Smartwatch (PADS) dataset on: https://physionet.org/content/parkinsons-disease-smartwatch/1.0.0/

## 9. Author Contributions

Seyed Mehdi Mohtavipour conceived the study, designed the methodology, implemented the software, conducted the experiments, analyzed the results, and wrote the manuscript.

## 10. Code Availability Statement

The source code developed for this study is publicly available at the following GitHub repository: https://github.com/MehdiMohtavipour/Latent-Motor-Abnormality-Scoring-of-Parkinson-s-Disease-

## 11. Ethics Statement

This study was conducted using the publicly available Parkinson’s Disease Smartwatch (PADS) dataset. The original PADS study was registered at ClinicalTrials.gov (NCT03638479) and was approved by the ethical board of the University of Münster and the physician’s chamber of Westphalia-Lippe (reference number: 2018-328-f-S). All participants provided written informed consent in the original study. The present work involved only secondary analysis of publicly available pseudonymized data, and no new participants were recruited.

